# The School-age Asthma Prognosis Score (SAPS): development and external validation in two European cohorts

**DOI:** 10.64898/2026.07.29.26359237

**Authors:** Ronny Makhoul, Myrofora Goutaki, Franco Romero, Mari Sasaki, Gesine Hansen, Pascal Heer, Matthias Volkmar Kopp, Philipp Latzin, Nicolas Regamey, Bianca Schaub, Elias Seidl, Ben D. Spycher, Claudia E. Kuehni

## Abstract

Prediction models for asthma remission in school-age are lacking, limiting clinicians’ ability to tailor follow-up. Most prediction tools focus on pre-school diagnosis or require lung function testing. We developed and validated a simple, history-based clinical prediction tool for asthma remission.

We analyzed prospective data from the Swiss Paediatric Airway Cohort (SPAC), including 1860 children (aged 5-16 years) with physician-diagnosed asthma. We derived asthma remission predictors from parental questionnaires capturing demographics, symptoms, triggers, and family history. We defined clinical remission at 2-3 years following asthma diagnosis, as absence of wheeze and inhaler use during the past 12 months. We developed the model using LASSO regression with multiple imputations for missing data, and assessed its performance by area under the curve (AUC), Hosmer-Lemeshow (HL) test, and calibration plots. We then derived a simplified score and validated it in the German All-Age Asthma Cohort (ALLIANCE).

From 12 candidate variables, the final score retained: sex, wheeze frequency, night-time awakening, exercise-induced wheeze, pollen-triggered wheeze, animal-triggered wheeze, maternal asthma, and paternal asthma. The score demonstrated moderate discrimination in the development cohort (AUC 0.71) and maintained discriminative ability in the external validation (AUC 0.71).

This practical, prognostic tool for asthma remission based only on clinical history, allows clinicians to identify children who have lower chances for remission, enabling their closer monitoring.

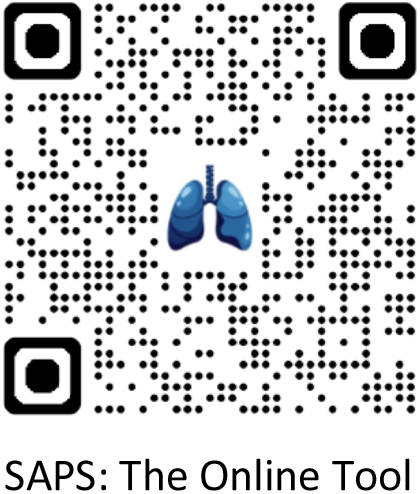

## Introduction

Asthma, one of the most common chronic diseases in childhood (1), is characterized by a highly heterogeneous natural history. While many children experience symptom resolution (‘remission’) during adolescence, others have persistent or worsening symptoms (2–4). When a child is diagnosed with asthma, parents often ask whether their child will outgrow it. Currently, clinicians rely largely on data from population-based studies to answer this question. Prognostic uncertainty can lead to anxiety for families and affect long-term management, potentially leading to overtreatment in children with a high chance of remission, or undertreatment in those with persistent disease (5–7).

Validated prognostic models for asthma remission or persistence for school-aged children are lacking. Previous tools, such as the Asthma Predictive Index (API), the Paediatric Asthma Risk Score (PARS), or the Predicting Asthma Risk in Children (PARC), have been designed for pre-schoolers and predict the development of asthma, not the prognosis of established asthma (7–11). The most notable exception is the prognostic model by Wang et al., derived from the Childhood Asthma Management Program (CAMP), which predicts asthma remission by early adulthood (12). However, this tool includes lung function tests, limiting its utility to settings where these are available. While spirometry and fractional exhaled nitric oxide (FeNO) are usually available in specialist care, their added value in predicting remission compared to a thorough clinical history remains debated (10, 12–15). A prediction tool that relies solely on standardized history-taking would be universally applicable.

We thus developed and externally validated a simple clinical risk score for asthma remission in school-aged children. We utilized data from the Swiss Paediatric Airway Cohort (SPAC) (16) for model development and the German All-Age Asthma Cohort (ALLIANCE) (17) for external validation. We hypothesized that a risk score based exclusively on clinical history could predict the probability of a child achieving clinical asthma remission over a 2-3 year period.

## Methods

### Study population and data sources

We analyzed longitudinal data from two large European clinical cohort studies. We developed the model using data from the Swiss Paediatric Airway Cohort (SPAC), a multicenter prospective cohort study integrated into routine care, which enrolls children referred to pediatric pulmonology clinics across Switzerland for common respiratory symptoms (16). Recruitment began in July 2017 and is ongoing. For external validation we used data from ALLIANCE, a multicenter prospective cohort recruiting patients with wheeze or asthma from respiratory outpatient clinics in Germany (17). Both studies are registered at ClinicalTrials.gov (SPAC: NCT03505216; ALLIANCE: NCT02496468). SPAC was approved by the Bern Cantonal Ethics Committee (Kantonale Ethikkomission Bern 2016-02176). The ALLIANCE study was approved by the relevant local ethics committees (17). Participants or their legal guardians provided written informed consent.

We included children aged 5-16 years with a physician-confirmed diagnosis of asthma at baseline, recruited to SPAC between July 2017 and January 2023, and to ALLIANCE between November 2013 and February 2022. In both cohorts, data were collected via comprehensive baseline and annual follow-up questionnaires to families capturing symptoms, medication use, and exposures. For the SPAC cohort, questionnaire data were supplemented by extraction of data from clinical records (referral reasons, diagnostic investigations, final diagnosis, and prescribed treatments), and were managed using a Research Electronic Data Capture (REDCap) database (18). Questions on asthma and allergic diseases in SPAC were adapted from the International Study of Asthma and Allergies in Childhood (ISAAC) (19) and the Leicester Respiratory Cohorts questionnaires (20). For ALLIANCE, data were derived from patient history (medical records), standardized questionnaires, structured interviews, and telephone interviews (17). We extracted the final analysis datasets in November 2025 (SPAC) and July 2025 (ALLIANCE).

### Outcome definition

We assessed asthma remission or persistence 2 to 3 years after the initial diagnosis, using data from the follow-up questionnaires from parents. When two questionnaires were available, we prioritized the third-year response to maximize the observation period. We defined asthma remission, as the absence of wheeze and no use of any asthma inhalers in the preceding 12 months (including short-acting beta agonists (SABA), inhaled corticosteroids (ICS), long-acting beta agonists (LABA), or combination therapy) (5, 11, 21). Participants not meeting these criteria were classified as having persistent asthma (reported wheeze, asthma inhaler use, or both, in the last 12 months).

### Candidate predictors

We selected candidate predictors of asthma remission through a rigorous, multi-step process to ensure clinical relevance, generalizability, and feasibility for real-world practice. Selection criteria included clinical importance, ease and reliability of measurement, cost, completeness of data, measurement precision, support from previous literature, suitability for diverse clinical settings, and ethical considerations. Following established guidance for prediction modelling (22–24), we prioritized variables that are readily available from patient history, clinical records, or standardized questionnaires, without requiring specialized testing. Based on these (3, 7, 10, 12–14, 25), and consensus discussions with pediatric pulmonologists, we identified 12 candidate predictors: sex, age, body mass index (BMI), hay fever, pollen-triggered wheeze, animal-triggered wheeze, wheeze frequency, exercise-induced wheeze, cough at night, night-time awakening by wheeze, paternal asthma, and maternal asthma. We excluded socioeconomic variables, as prior evidence showed no incremental predictive value and limited generalizability (11, 26). We calculated BMI z-scores using the 2007 WHO growth reference (27). We tested continuous predictors (age and BMI) for departures from linearity using logistic regression models with linear, quadratic, restricted cubic splines, and piecewise linear terms. We retained the linear form when these best fit the data (22, 28).

### Handling of missing data

In the development dataset, we compared baseline characteristics of included participants and those lost to follow-up to assess potential selection bias. Where reasonable, we recoded missingness to zero, indicating the absence of that symptom or treatment (**Supplementary Methods**). For all other missing outcome or predictor variables, we used multiple imputations by chained equations (MICE), applying a random-forest algorithm with 50 imputed datasets (24, 29, 30). The imputation model included all candidate predictors, relevant demographic variables, and auxiliary variables (**Supplementary Table E1** and **E2** for the full list of auxiliaries). In the external validation dataset, where missingness was minimal (0-2% across predictors), we set missing values to zero (**Supplementary Table E4**).

### Sample size and model complexity

According to recommendations (22, 31). The SPAC dataset was sufficient to support development of a model with up to 13 parameters while minimizing the risk of overfitting. We provide detailed calculations and underlying assumptions in **Supplementary Methods**.

### Model development and validation

For model development, we applied least absolute shrinkage and selection operator (LASSO)- penalized logistic regression on 50 multiply imputed datasets, retaining predictors that had non-zero coefficients in ≥90% of imputations and pooling coefficients, including zeros (22, 24, 29, 32, 33). We summarized the apparent performance by discrimination (area under the receiver operating characteristic curve (AUC)), calibration (plots and Hosmer-Lemeshow (HL)), and overall accuracy (Brier score). We internally validated with 600 bootstrap resamples with nested imputation and out-of-bag testing (22, 24). Model development and internal validation framework is illustrated in **Supplementary Figure E1**. We derived scores by multiplying pooled coefficients by a scaling factor (10, 5, or 3), rounding the results, and recalibrating using logistic regression with total points as a linear predictor. We then calculated the predicted risk for each point total across the candidate scaled scores. We selected the final scoring that provided the optimal balance between simplicity and predictive performance (22).

We externally validated the final risk score in the independent ALLIANCE cohort. We harmonized predictors and outcome to match the SPAC definitions (17) to the extent permitted by the data (**Table 1**, **Supplementary Tables E3**, and **E4**). We applied the score ‘as is’ (strict validation), without refitting or updating the coefficients. We determined the model’s generalizability to the new population by evaluating discrimination (AUC), calibration (calibration plots and HL test), and overall accuracy (Brier score) (22). The comprehensive methodological framework for model development and validation and the TRIPOD checklist (23) are detailed in **Supplementary Appendix**.

**Table 1:** Baseline characteristics of children with available follow-up information in the development cohort (SPAC, N=1237), and the validation cohort (ALLIANCE, N=305) of the School-age Asthma Prognosis Score (SAPS)

|  | SPAC<br>N (%) | ALLIANCE<br>N (%) |
| --- | --- | --- |
| **Participants** | 1237 (100) | 305 (100) |
| *Individual characteristic* |  |  |
| **Sex, male** | 757 (61) | 199 (65) |
| **Age, years, median (IQR)** | 10 (8-13) | 9 (7-12) |
| **Body mass index (BMI), median (IQR)** | 17 (16-19) | 18 (16-21) |
| Missing weight, height, or both | 9 (1) | 0 (0) |
| *Clinical symptom severity (last 12 months)* |  |  |
| **Wheeze frequency, episodes\*\*** |  |  |
| None | 461 (37) | 79 (26) |
| SPAC: 1-3, ALLIANCE: 1-2 | 415 (34) | 79 (26) |
| SPAC: $\geq 4$, ALLIANCE: $\geq 3$ | 345 (28) | 147 (48) |
| Missing data | 16 (1) | 0 (0) |
| **Exercise-induced wheeze, yes** | 579 (47) | 185 (61) |
| No | 644 (52) | 120 (39) |
| Missing data | 14 (1) | 0 (0) |
| **Night-time awakening by wheeze, yes** | 427 (35) | 138 (45) |
| No | 790 (63) | 167 (55) |
| Missing data | 20 (2) | 0 (0) |
| *Comorbidities* |  |  |
| **Hay fever, yes** | 615 (50) | 130 (43) |
| No | 603 (48) | 175 (57) |
| Missing data | 19 (2) | 0 (0) |
| *Allergic triggers* |  |  |
| **Pollen-triggered wheeze, yes** | 440 (36) | 123 (40) |
| No | 797 (64) | 182 (60) |
| Missing data* | 0 (0) | 0 (0) |
| **Animal-triggered wheeze, yes** | 233 (19) | 94 (31) |
| No | 1004 (81) | 211 (69) |
| Missing data* | 0 (0) | 0 (0) |
| *Family history* |  |  |
| **Maternal asthma, yes** | 266 (22) | 72 (24) |
| No | 971 (78) | 233 (76) |
| Missing data | (0) | 0 (0) |
| **Paternal asthma, yes** | 215 (17) | 64 (21) |
| No | 1022 (83) | 241 (79) |
| Missing data | (0) | 0 (0) |

IQR: interquartile range; \*Missingness for ALLIANCE and specific SPAC predictors was set to zero (See 'Handling of missing data' in Methods); \*\*We harmonized the wheeze frequency variable categories, as they differ in SPAC and ALLIANCE data.

### Software and reproducibility

We conducted our analyses in Python (**Supplementary Table E5**). We performed sample size calculations in R using the pmsampsize package (31). We ensured reproducibility by fixing the random seed for all non-deterministic operations.

## Results

We included 1860 children in the development cohort (SPAC), of whom 1237 (67%) completed a second- or third-year follow-up, while 623 (33%) were lost by the time of follow-up. Asthma had remitted in 214 (17%) (**Supplementary Figure E2** for the study flow diagram). Among the 1237 children in the development cohort, 757 (61%) were boys; median age was 10.2 years (IQR 8-13). Most parents had at least intermediate education (92% of mothers, 91% of fathers), and were employed (80%, 94%) (**Supplementary Table E1**). Baseline characteristics of participants with and without follow-up were comparable, with minor differences in socio-economic factors and BMI (**Supplementary Table E1**).

In the external validation cohort (ALLIANCE), we included 305 children aged 5-16 years at baseline. Of those, 66 (22%) had asthma remission within 2-3 years. Baseline characteristics were broadly similar (**Table 1**).

### Model development and internal validation

Of the 12 candidate predictors entered into the LASSO regression, 10 were retained in the final model. BMI and cough at night were excluded due to weak predictive value (zero coefficients in >10% of imputations). The retained predictors included: sex, age, hay fever, pollen-triggered wheeze, animal-triggered wheeze, wheeze frequency, exercise-induced wheeze, night-time awakening by wheeze, paternal asthma, and maternal asthma. The model coefficients are presented in **Supplementary Table E6**.

The model showed moderate apparent discrimination, with an AUC of 0.71 (**Supplementary Figure E3-A**). The model was well-calibrated (HL p = 0.48), with close agreement between predicted and observed risks in the calibration plots (**Supplementary Figure E3-B**). In internal validation using bootstrapping, the optimism-corrected discrimination and calibration was somewhat lower (AUC 0.68, and HL p = 0.1) (**Supplementary Figure E4**).

### Points-based risk score: the School-age Asthma Prognosis Score (SAPS)

Comparisons of scaling factors (multiplying coefficients by 10, 5, or 3) revealed that a factor of 3 offered the best balance between clinical usability and predictive performance (**Supplementary Table E7** and **E8**). This simplified model retained comparable predictive performance to the full development model (AUC = 0.70, HL p = 0.68, Brier = 0.14), see ROC curve and calibration plots (**Figure 1A**, and **1B**). Rounding coefficients to the nearest integer resulted in age and hay fever receiving zero points, excluding them from the simplified tool. The final score therefore comprised a parsimonious 8 predictors (six questions), all of which were associated with a reduced probability of remission (negative points); with a base score of 11 points. The strongest predictors of no remission were wheeze frequency of 4 episodes or more (-2 points), night-time awakening by wheeze (-2 points), and pollen-triggered wheeze (-2 points). To calculate a patent’s total score, subtract the assigned points for each question from 11 (**Figure 2**). The corresponding score-to-risk curve illustrates the estimated predicted probability of asthma remission 2-3 years later for all scores. To facilitate clinical implementation, the model is available as the School-age Asthma Prognosis Score (SAPS), a user-friendly, web-based calculator (https://saps.predmod.dev).

**Figure 1:**
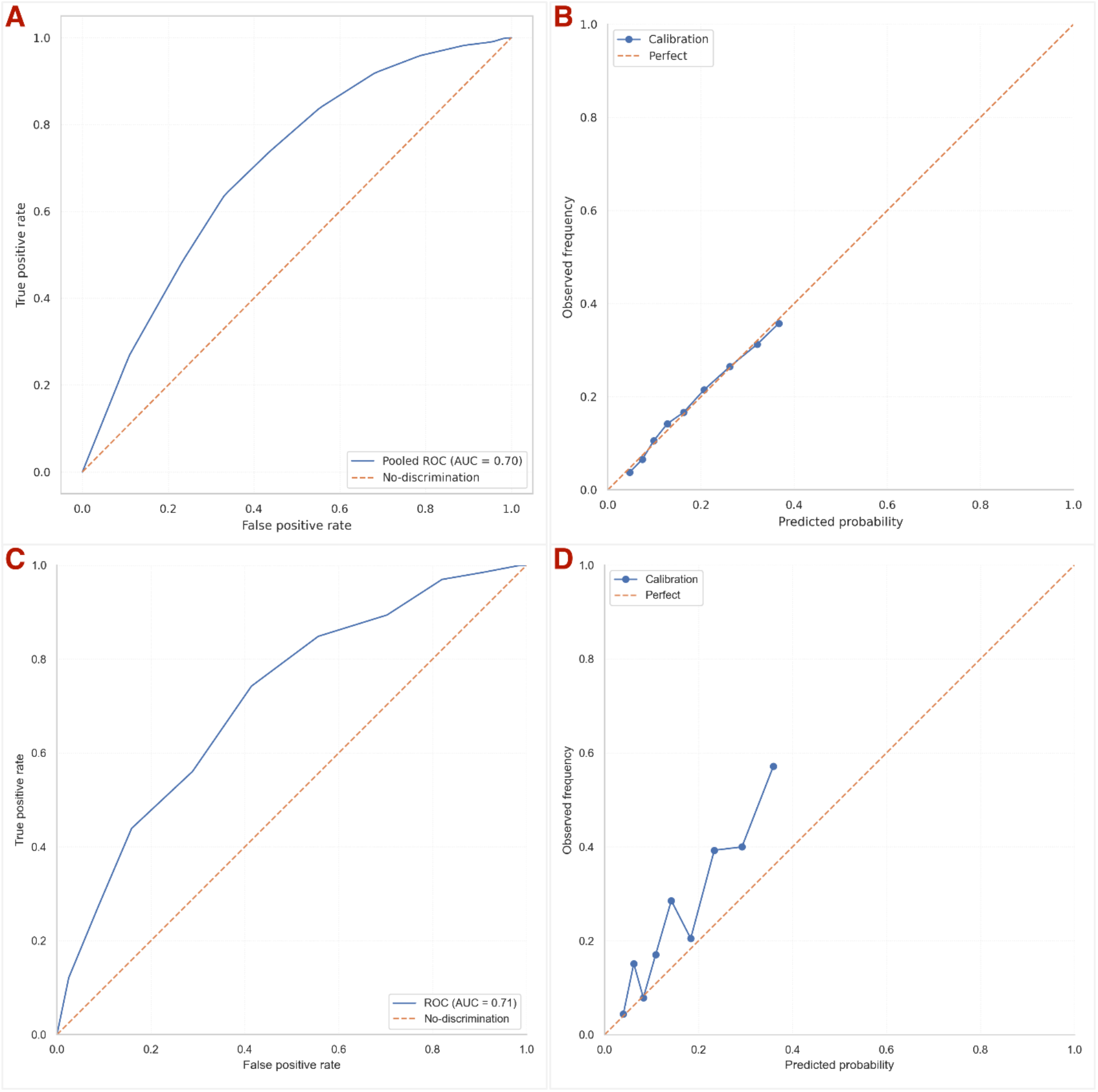
Pooled ROC curve and calibration plots (development (A, B) versus external validation (C, D)) of the asthma remission points-based risk score.

**Figure 2:**
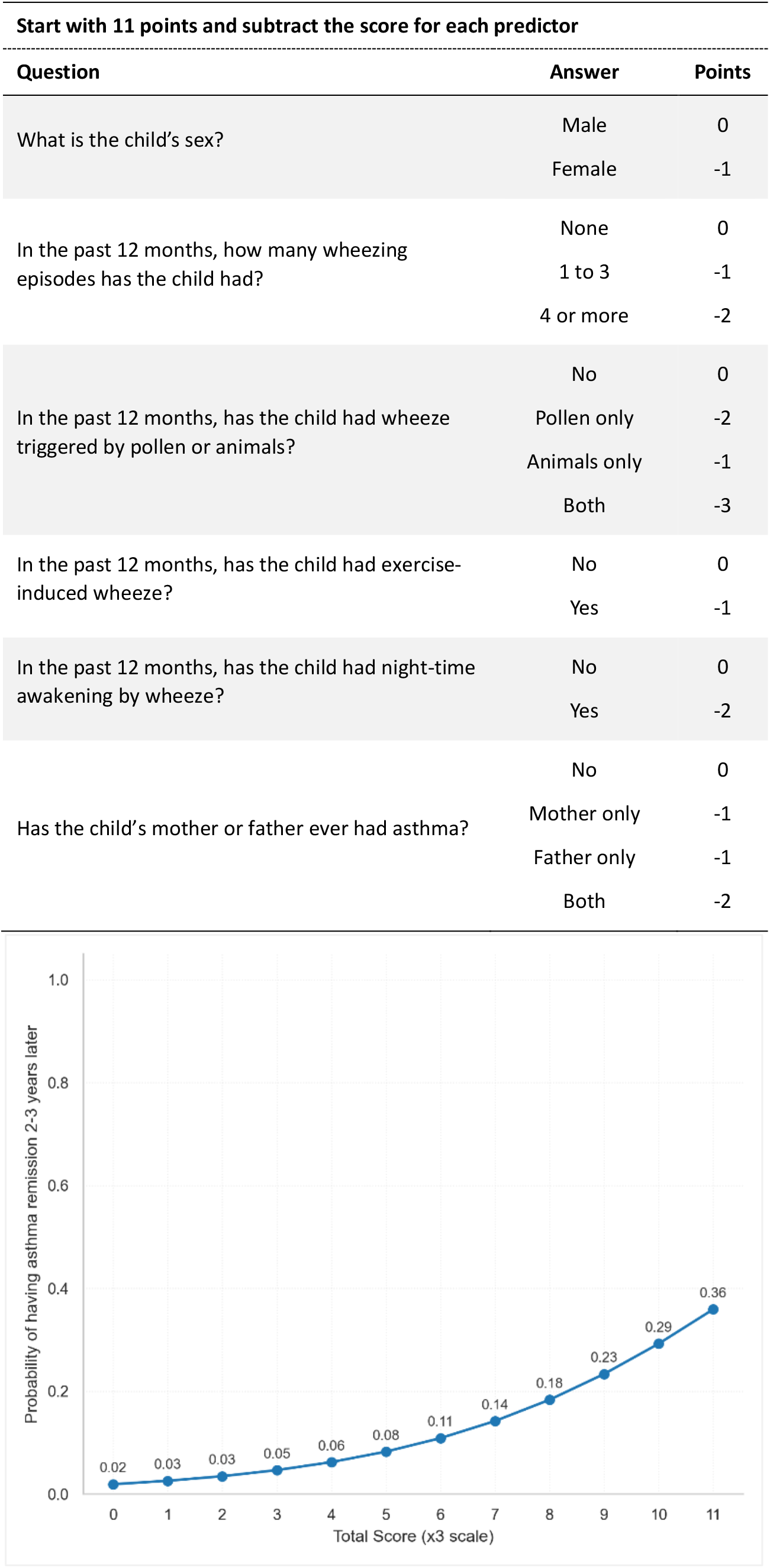
Calculation of the School-age Asthma Prognosis Score (SAPS) and the estimated probability of medium-term asthma remission (2-3 years) in children aged 5-16 years based on total score.

### External validation

In the ALLIANCE cohort, the score demonstrated fair transportability (**Table 2**, **Figure 1**). Discrimination was maintained, with an AUC of 0.71. However, calibration deteriorated (p=0.003), reflecting the discrepancy between predicted probabilities and observed frequency. The ROC curve and calibration plots (**Figure 1C**, and **1D**) illustrate this performance.

**Table 2:** Overview of performance metrics of the clinical prediction model for asthma remission in the development (SPAC) and validation (ALLIANCE) cohorts

|  | Brier score | AUC | HL p-value |
| --- | --- | --- | --- |
| Apparent performance | 0.137 | 0.706 | 0.484 |
| Internal validation | 0.136* | 0.678* | 0.113 |
| Point-based score | 0.138 | 0.701 | 0.679 |
| External validation<sup>a</sup> | 0.160 | 0.713 | 0.003 |

\*Optimism-corrected estimate; <sup>a</sup>External validation using ALLIANCE data.

## Discussion

The School-age Asthma Prognosis Score (SAPS) is an externally validated clinical prediction tool to estimate the probability of asthma remission in school-aged children. We developed it using data of children from a clinical setting (respiratory outpatient clinics) and includes eight readily available items from patient history (wheeze severity and triggers, sex, and parental respiratory history). Our model demonstrated stable moderate discrimination in both internal and external validation. However, calibration deteriorated in the new population, reflecting differences in asthma prevalence at follow-up. This suggests that while the predictor set is robust, the model requires recalibration to support transportability.

Prior asthma prediction tools had focused on prediction of asthma incidence. Several tools described the development of asthma in pre-school children (e.g., API, PARS, or PARC) (8, 9, 11). In contrast, tools predicting remission in school-aged children with established asthma are lacking (13), with the exception of the School-Age Asthma Remission model, developed from the Childhood Asthma Management Program (CAMP) (12). While the CAMP model achieved higher discrimination (AUC 0.79- 0.85), it requires spirometry (FEV1/FVC), methacholine challenge testing, and specific IgE measurements. We developed a simpler model to create a history-only tool. While our approach accepts a trade-off in discrimination, providing a 70% probability of correctly distinguishing between a child with persistent disease and one who achieves remission (AUC 0.70), it eliminates the need for specialized diagnostics, ensuring the tool is universally applicable, in resource-limited settings where objective testing is often inaccessible.

We observed a predicted probability range of 2% to 36%. While the model does not identify a group with a high (e.g. >80%) chance of remission, it effectively stratifies risk: a child with the maximum score of 11, is 18 times more likely to remit than one with the lowest. Clinically, this makes the model particularly powerful as a ‘rule-out’ tool for remission; a low score (e.g., 2% probability) provides strong evidence for asthma persistence, supporting the decision to continue maintenance therapy.

Our predictors align with established prognostic factors described in foundational epidemiological studies (2, 34–37). We identified wheeze frequency of 4 episodes or more, night-time awakening by wheeze, and pollen-triggered wheeze as the strongest clinical predictors of asthma persistence. The prognostic weight of wheeze frequency and night symptoms aligns with the prognostic relevance of clinical severity described in cohort studies (34, 35, 37), where indicators of symptom severity in childhood strongly predicted adult persistence. The impact of allergic triggers (pollen) confirms the link between atopy and asthma persistence, mirroring findings described in literature and recent work in the SPAC cohort (2, 25). While male sex is a well-recognized risk factor for asthma incidence in early childhood (36), we found it a predictor of remission in school age, consistent with literature on changes in hormonal status and airway geometry occurring during puberty (38, 39).

A key strength of our study is the use of multicenter, prospective data from large clinical cohorts with standardized follow-up (SPAC and ALLIANCE) rather than general population samples. Hospital-based study populations ensure the tool is derived from and applicable to patients encountered in pediatric clinical practice rather than to mainly healthy children as in birth cohorts. While external validation in a similar setting confirmed the model’s discriminative ability, its transportability and use in other healthcare settings requires further validation. The observed deterioration in calibration was likely driven by the higher remission rate in the validation set (22%) compared with the development set (17%). This mismatch typically causes ‘miscalibration-in-the-large’, affecting the baseline risk estimate while leaving discrimination intact (22). Consequently, local recalibration may be required to optimize absolute risk estimates before implementation in new settings. We adhered to TRIPOD reporting guidelines (23) and employed contemporary methods for model development, including multiple imputation, and LASSO penalization to minimize the overfitting (5, 24). Furthermore, by strictly focusing on readily available clinical variables, we prioritized model parsimony. This ensures the tool remains practical for settings where complex diagnostics are inaccessible.

We acknowledge several other limitations. SPAC and ALLIANCE cohorts represent a predominantly tertiary care population rather than primary care, comprised of children with mild-to-moderate asthma. We defined asthma remission using self-reported symptoms, which are subject to recall bias and misinterpretation. However, our predictors were derived from standardized instruments, adapted from the ISAAC and Leicester Respiratory Cohorts questionnaires (19, 20). The sample size of the external validation cohort (ALLIANCE) was relatively small which may limit the precision of our performance estimates. Our definition of asthma remission represents clinical symptoms remission rather than biological or complete remission. Our 2-3 year outcome window may be insufficient to describe long-term remission.

In practice, our tool allows clinicians to identify children with a low probability of remission and prioritize them for closer monitoring, while families of children with a high probability of remission can be offered hope for improvement. Looking forward, future research should explore the integration of objective measurements into this history-only tool. For the tertiary care setting, potential extensions could include spirometry and FeNO (primary care), allergic sensitization profiles (secondary care) and airway hyperresponsiveness or body plethysmography. The gain in predictive performance of such measurements would need to be weighed against their added complexity and cost. Additionally, dynamic prediction approaches incorporating repeated measures over time should be tested to determine whether updating predictions enhances accuracy and clinical utility (22, 40). In conclusion, our externally validated clinical prediction model provides a simple, accessible tool for assessing long-term asthma prognosis in school-aged children, enabling practical risk estimation at the point of care.

## Declaration of interests

Claudia E. Kuehni, Myrofora Goutaki, Ronny Makhoul, Franco Romero, Mari Sasaki, Ben D. Spycher, Nicolas Regamey, Pascal Heer, and Elias Seidl have no conflicts of interest.

Philipp Latzin declares grants or contracts from Vertex and OM Pharma; Consulting fees from IQone; Payment or honoraria for lectures, presentations, speakers bureaus, manuscript writing or educational events from Vertex, Vifor, OM Pharma, Astra Zeneca; Participation on a Data Safety Monitoring Board or Advisory Board of Allecra, Santhera, Vertex, OM Pharma.

Gesine Hansen declares consulting fees from Sanofi; Payment or honoraria for lectures, presentations, speakers bureaus, manuscript writing or educational events from MedUpdate, Abbvie, Springer Verlag.

Matthias V. Kopp declares payment or honoraria for lectures, presentations, speakers bureaus, manuscript writing or educational events from Infectopharm GmbH and Allergopharma GmbH; Participation on a Data Safety Monitoring Board or Advisory Board of Allergopharma GmbH; Past President – President of the Society of Paediatric Pulmonology (till 2025).

Bianca Schaub declare grants or contracts from DFG, BMFTR, and EU; Consulting fees from GlaxoSmithKline, Novartis, Sanofi, and Astra Zeneca; Payment or honoraria for lectures, presentations, speakers bureaus, manuscript writing or educational events from Sanofi and Astra Zeneca; Support for attending meetings and/or travel from Astra Zeneca; Participation on a Data Safety Monitoring Board or Advisory Board of Sanofi and Astra Zeneca; Leadership or fiduciary role in other board, society, committee or advocacy group, paid or unpaid of DGAKI, EAACI, ERS.

All declarations of all authors are outside the submitted work.

## Abbreviations used

ALLIANCE: German All-Age Asthma Cohort
API: Asthma Predictive Index
AUC: area under the curve
BMI: body mass index
CAMP: Childhood Asthma Management Program
FeNO: Fractional exhaled nitric oxide
FEV1: Forced expiratory volume in 1 second
FVC: Forced vital capacity
HL: Hosmer-Lemeshow
ICS: Inhaled corticosteroids
IQR: Interquartile range
ISAAC: International Study of Asthma and Allergies in Childhood
LABA: Long-acting beta agonists
LASSO: Least absolute shrinkage and selection operator
MICE: Multiple imputations by chained equations
PARC: Predicting Asthma Risk in Children
PARS: Paediatric Asthma Risk Score
REDCap: Research Electronic Data Capture
ROC: Receiver operating characteristic
SABA: Short-acting beta agonists
SAPS: School-age Asthma Prognosis Score
SPAC: Swiss Paediatric Airway Cohort
WHO: World Health Organization

## Acknowledgments

We thank the families who took part in SPAC and ALLIANCE studies. We thank the members of the SPAC Study team and the ALLIANCE Study team. Members of the current and past SPAC Study team are: T. Schürmann and C. Bieli (Cantonal Hospital Aarau, Aarau, Switzerland); A. Jochmann, D. Trachsel and J. Usemann (University Children’s Hospital Basel, Basel, Switzerland); P. Latzin, C. Casaulta, M. Bullo, I. Korten, E. Kieninger, B. Frauchiger, B. Vomsattel, S. Yammine and C.C.M de Jong (Inselspital, Bern University Hospital, University of Bern, Bern, Switzerland); P. Iseli (Cantonal Hospital Graubünden, Chur, Switzerland); K. Hoyler (private paediatric pulmonologist, Horgen, Switzerland); S. Blanchon, S. Guerin, I. Rochat and C. Fernandez-Elviro (Lausanne University Hospital, University of Lausanne, Lausanne); N. Regamey, M. Lurà, M. Hitzler, K. Hrup, E. Sidler, P. Heer and C. Bernold (Children’s Hospital of Central Switzerland, Lucerne, Switzerland); J. Barben (Children’s Hospital St. Gallen, St. Gallen, Switzerland); O. Sutter and L. Krüger (private paediatric practice, Worb, Bern, Switzerland); A. Moeller, E. Seidl, G. Signorelli, YT. Lam, G. Buggle, S. Beck and L. Benz (University Children’s Hospital Zurich, Zurich, Switzerland); and C.E. Kuehni, M. Goutaki, C. Ardura-Garcia, D. Berger, S. Glick, B. Guerra, T. Krasnova, M.C. Mallet, E. Pedersen, F. Romero, M. Sasaki, V. Weihrauch, and R. Makhoul (Institute of Social and Preventive Medicine, University of Bern, Bern, Switzerland). We thank T. Krasnova, M.C. Mallet, B. Guerra, G. Ly, M. Ryser, M. Frei, E. Schneider, S. Chellakudam, and L. Z. Volkart (University of Bern, Bern, Switzerland) for their contributions in the acquisition and entering of data.

## Declaration of Generative AI and AI-assisted technologies in the writing process

During the preparation of this work the authors used Google AI to improve readability and language. After using this tool, the authors reviewed and edited the content as needed and takes full responsibility for the content of the publication.

## Contributors

RM drafted the manuscript. MG, BDS, and CEK made substantial contributions to the conception, design, and interpretation of this work. RM and FR made contributions to the acquisition and entering of data. RM verified the underlying data and performed statistical analyses. All authors made contributions to the interpretation of the data for this work and had final responsibility for the decision to submit for publication.

## Data sharing

The SPAC and ALLIANCE study protocols, consent forms, definitions, and derivation of clinical characteristics and outcomes, regulatory documents, information about requests for data access, and other relevant study materials are available online (www.spac-study.ch and www.dzl.de). SPAC study data is not deposited at an open access repository, as participants were not asked to give consent to have their data deposited publicly. Requests for partial datasets for specific analyses including individual patient data and a data dictionary defining each included field can be addressed to Prof Kuehni upon reasonable request. Other data (analysis documentation and Python code) can be made available on reasonable request to the corresponding author.

