## Supplementary materials for "The School-age Asthma Prognosis Score (SAPS): development and external validation in two European cohorts"

**Table E1:** Baseline characteristics and suggested candidate predictors of the School-age Asthma Prognosis Score (SAPS), comparing children who completed 2-3 years follow-up with those who were lost to follow-up (N=1860)

| | **Follow-up available** N (%) 1237 (67) | **Lost to follow-up** N (%) 623 (33) | ***p-value*** * |
|---|---|---|---|
| *Individual characteristic* | | | |
| **Sex**, male | 757 (61) | 374 (60) | 0.663 |
| **Age** at baseline, years, median (IQR) | 10 (8-13) | 10 (8-13) | 0.796 |
| **Body mass index (BMI)** at baseline, median (IQR) | 17 (16-19) | 18 (16-21) | 0.004 |
| Missing weight, height, or both | 9 (1) | 9 (1) | |
| *Clinical symptom severity* | | | |
| **Wheeze frequency**, episodes | | | |
| None | 461 (37) | 244 (39) | 0.788 |
| 1-3 | 415 (34) | 201 (32) | |
| 4-12 | 257 (21) | 120 (19) | |
| > 12 | 88 (7) | 45 (7) | |
| Missing data | 16 (1) | 13 (2) | |
| **Exercise-induced wheeze**, yes | 579 (47) | 270 (43) | 0.198 |
| No | 644 (52) | 343 (55) | |
| Missing data | 14 (1) | 10 (2) | |
| **Night-time awakening by wheeze**, night(s) per week | | | |
| Never | 790 (64) | 400 (64) | 0.940 |
| < 1 | 275 (22) | 134 (22) | |
| ≥ 1 | 152 (12) | 78 (13) | |
| Missing data | 20 (2) | 11 (2) | |
| **Cough at night**, yes | 533 (43) | 271 (44) | 0.778 |
| No | 673 (54) | 331 (53) | |
| Missing data | 31 (3) | 21 (3) | |
| *Comorbidities* | | | |
| **Hay fever**, yes | 615 (50) | 280 (45) | 0.060 |
| No | 380 (31) | 194 (31) | |
| Do not know | 223 (18) | 138 (22) | |
| Missing data | 19 (2) | 11 (2) | |
| *Allergic triggers* | | | |
| **Pollen-triggered wheeze** | | | |
| Never | 548 (44) | 277 (45) | 0.035 |
| Sometimes | 265 (21) | 133 (21) | |
| Often | 175 (14) | 58 (9) | |
| Missing data | 249 (20) | 155 (25) | |
| **Animal-triggered wheeze** | | | |
| Never | 738 (60) | 367 (59) | 0.112 |
| Sometimes | 128 (10) | 63 (10) | |
| Often | 105 (9) | 34 (6) | |
| Missing data | 266 (22) | 159 (26) | |

| | Follow-up available N (%) 1237 (67) | Lost to follow-up N (%) 623 (33) | *p-value* * |
|---|---|---|---|
| *Family history* | | | |
| **Maternal asthma**, yes | 266 (22) | 124 (20) | 0.459 |
| No | 971 (78) | 499 (80) | |
| **Paternal asthma**, yes | 215 (17) | 105 (17) | 0.826 |
| No | 1022 (83) | 518 (83) | |
| *Socio-economic factors* | | | |
| **Education level**, mother | | | |
| Basic | 78 (6) | 61 (10) | 0.006 |
| Intermediate | 671 (54) | 339 (54) | |
| Higher | 464 (38) | 200 (32) | |
| Missing data | 24 (2) | 23 (4) | |
| **Education level**, father | | | |
| Basic | 74 (6) | 54 (9) | <0.001 |
| Intermediate | 542 (44) | 302 (49) | |
| Higher | 582 (47) | 235 (38) | |
| Missing data | 39 (3) | 32 (5) | |
| **Employment status**, mother | | | |
| Unemployed | 11 (1) | 7 (1) | 0.860 |
| Employed | 984 (80) | 481 (77) | |
| Other (housewife, training, etc) | 212 (17) | 105 (17) | |
| Missing data | 30 (2) | 30 (5) | |
| **Employment status**, father | | | |
| Unemployed | 13 (1) | 4 (1) | 0.484 |
| Employed | 1158 (94) | 560 (90) | |
| Other (‘houseman’, training, etc) | 16 (1) | 11 (2) | |
| Missing data | 50 (4) | 48 (8) | |

IQR: interquartile range; *p-values were calculated using the Chi-square test, except for age and BMI, the Mann-Whitney U test was used.

**Table E2:** Auxiliary variables[a] used in the multiple imputation in the development cohort (SPAC) of the clinical prediction model for asthma remission

| **Variable** |
|---|
| *Candidate predictors* |
| Cough without cold |
| ICS at baseline |
| Rapid relief inhaler |
| Maternal hay fever |
| Paternal hay fever |
| Interference with daily activity by wheeze |
| Cough more than others |
| *Demographic factors* |
| Education level (mother) |
| Education level (father) |
| Employment status (mother) |
| Employment status (father) |
| Lives in Switzerland since birth (child) |
| Lives in Switzerland since birth (mother) |
| Lives in Switzerland since birth (father) |
| Swiss citizen (child) |
| Swiss citizen (mother) |
| Swiss citizen (father) |
| Child was breastfed |

[a] Candidate predictors which were eventually set aside from using as a predictor in the model, and demographic factors.

**Table E3:** Baseline characteristics of included children in the development cohort (SPAC) by remission status (N=1237)

| | Asthma 2-3 years later | | |
|---|---|---|---|
| | **Remission** N (%) 214 (17) | **Persistence** N (%) 1023 (83) | **Total** N (%) 1237 (100) |
| *Individual characteristic (at baseline)* | | | |
| **Sex**, male | 150 (70) | 607 (59) | 757 (61) |
| **Age** at baseline, years, median (IQR) | 10 (8-13) | 10 (8-13) | 10 (8-13) |
| **Body mass index (BMI)** at baseline, median (IQR) | 17 (16-19) | 17 (16-19) | 17 (16-19) |
| Missing weight, height, or both | 2 (1) | 7 (1) | 9 (1) |
| *Clinical symptom severity (in last 12 months at baseline)* | | | |
| **Wheeze without cold**, yes | 42 (20) | 346 (34) | 388 (31) |
| No | 141 (66) | 478 (47) | 619 (50) |
| Missing data | 31 (15) | 199 (20) | 230 (19) |
| **Wheeze frequency**, episodes | | | |
| None | 118 (55) | 343 (34) | 461 (37) |
| 1-3 | 64 (30) | 351 (34) | 415 (34) |
| 4-12 | 21 (10) | 236 (23) | 257 (21) |
| > 12 | 7 (3) | 81 (8) | 88 (7) |
| Missing data | 4 (2) | 12 (1) | 16 (1) |
| **Exercise-induced wheeze**, yes | 65 (30) | 514 (50) | 579 (47) |
| No | 148 (69) | 496 (49) | 644 (52) |
| Missing data | 1 (1) | 13 (1) | 14 (1) |
| **Night-time awakening by wheeze**, night(s) per week | | | |
| Never | 173 (81) | 617 (60) | 790 (63) |
| < 1 | 22 (10) | 253 (25) | 275 (22) |
| ≥ 1 | 16 (8) | 136 (13) | 152 (12) |
| Missing data | 3 (1) | 17 (2) | 20 (2) |
| *Comorbidities* | | | |
| **Hay fever**, yes | 89 (42) | 526 (51) | 615 (50) |
| No | 84 (39) | 296 (29) | 380 (31) |
| Do not know | 37 (17) | 186 (18) | 223 (18) |
| Missing data | 4 (2) | 15 (2) | 19 (2) |
| *Allergic triggers* | | | |
| **Pollen-triggered wheeze** | | | |
| Never | 120 (56) | 428 (42) | 548 (44) |
| Sometimes | 24 (11) | 241 (24) | 265 (21) |
| Often | 18 (8) | 157 (15) | 175 (14) |
| Missing data | 52 (24) | 197 (19) | 249 (20) |
| **Animal-triggered wheeze** | | | |
| Never | 134 (63) | 604 (59) | 738 (60) |
| Sometimes | 11 (5) | 117 (11) | 128 (10) |
| Often | 7 (3) | 98 (10) | 105 (9) |
| Missing data | 62 (29) | 204 (20) | 266 (22) |
| *Family history* | | | |
| **Maternal asthma**, yes | 34 (16) | 232 (23) | 266 (22) |
| No | 180 (84) | 791 (77) | 971 (79) |
| **Paternal asthma**, yes | 26 (12) | 189 (19) | 215 (17) |
| No | 188 (88) | 834 (82) | 1022 (83) |

IQR: interquartile range.

**Table E4:** Baseline characteristics of included children in the validation cohort (ALLIANCE) by remission status (N=305)

| | Asthma 2-3 years later | | |
|---|---|---|---|
| | **Remission** N (%) 66 (22) | **Persistence** N (%) 239 (78) | **Total** N (%) 305 (100) |
| *Individual characteristic (at baseline)* | | | |
| **Sex**, male | 49 (74) | 150 (63) | 199 (65) |
| **Age**, years, median (IQR) | 9 (7-12) | 9 (7-12) | 9 (7-12) |
| **Body mass index (BMI)**, median (IQR) | 17 (16-20) | 18 (16-21) | 18 (16-21) |
| Missing weight, height, or both | 0 (0) | 0 (0) | 0 (0) |
| *Clinical symptom severity (in last 12 months at baseline)* | | | |
| **Wheeze frequency**, episodes | | | |
| None | 28 (42) | 50 (21) | 78 (26) |
| 1 | 6 (9) | 26 (11) | 32 (10) |
| 2 | 18 (27) | 29 (12) | 47 (15) |
| 3-4 | 10 (15) | 62 (26) | 72 (24) |
| > 4 | 4 (6) | 71 (30) | 75 (25) |
| Missing data | 0 (0) | 1 (<1) | 1 (<1) |
| **Exercise-induced wheeze**, yes | 28 (42) | 157 (66) | 185 (61) |
| No | 38 (58) | 80 (34) | 118 (39) |
| Missing data | 0 (0) | 2 (1) | 2 (1) |
| **Night-time awakening by wheeze**, yes | 22 (33) | 116 (49) | 138 (45) |
| No | 44 (67) | 122 (51) | 166 (54) |
| Missing data | 0 (0) | 1 (<1) | 1 (<1) |
| **Hay fever** *(ever)*, yes | 22 (33) | 108 (45) | 130 (43) |
| No | 43 (65) | 130 (54) | 173 (57) |
| Missing data | 1 (2) | 1 (<1) | 2 (1) |
| *Allergic triggers (ever)* | | | |
| **Pollen-triggered wheeze**, yes | 22 (33) | 101 (42) | 123 (40) |
| No | 44 (67) | 136 (57) | 180 (59) |
| Missing data | 0 (0) | 2 (1) | 2 (1) |
| **Animal-triggered wheeze**, yes | 9 (14) | 85 (36) | 94 (31) |
| No | 57 (86) | 152 (64) | 209 (69) |
| Missing data | 0 (0) | 2 (1) | 2 (1) |
| *Family history* | | | |
| **Maternal asthma**, yes | 4 (6) | 68 (29) | 72 (24) |
| No | 60 (91) | 169 (71) | 229 (75) |
| Missing data | 2 (3) | 2 (1) | 4 (1) |
| **Paternal asthma**, yes | 11 (17) | 53 (22) | 64 (21) |
| No | 53 (80) | 184 (77) | 237 (78) |
| Missing data | 2 (3) | 2 (1) | 4 (1) |

IQR: interquartile range; Structural missing data were re-coded to zero '0'.

**Table E5:** Software environment and Python library versions used for the data analysis

| Title | Version |
|---|---|
| python | 3.12.3 |
| numpy | 1.26.4 |
| pandas | 2.3.1 |
| matplotlib | 3.9.2 |
| scipy | 1.15.3 |
| scikit-learn | 1.7.0 |
| miceforest | 5.7.0 |
| pmsampsize (in R) | 1.1.3 (4.4.3) |

**Table E6:** Predictor selection and pooled coefficients for the School-age Asthma Prognosis Score (SAPS) in the development cohort (SPAC)

| Variable | Selection | Coefficient |
|---|---|---|
| Female sex | 50/50 | -0.261 |
| Age | 49/50 | -0.052 |
| BMI (z-score) | 39/50 (Less than 45) | Dropped out |
| Pollen-triggered wheeze | 50/50 | -0.506 |
| Animal-triggered wheeze | 50/50 | -0.373 |
| Hay fever | 47/50 | -0.129 |
| Wheeze frequency 1 (1-3) | 50/50 | -0.241 |
| Wheeze frequency 2 (≥ 4) | 50/50 | -0.304 |
| Exercise-induced wheeze | 49/50 | -0.198 |
| Night-time awakening by wheeze | 50/50 | -0.755 |
| Cough at night | 14/50 (Less than 45) | Dropped out |
| Paternal asthma | 50/50 | -0.233 |
| Maternal asthma | 50/50 | -0.221 |

Intercept (average) = -0.609; **Selection refers to non-zero value in at least 90% of imputations**; Age and BMI are continuous variables; Wheeze frequency is categorical **ordinal** variable (cumulative); Rest of variables are binary.

**Table E7:** Derivation of the School-age Asthma Prognosis Score (SAPS): Comparison of candidate point-based models for asthma remission using scaling factors of 10, 5, and 3

| Variable | x 10 | x 5 | x 3 |
|---|---|---|---|
| **Base score (total points)** | **33** | **17** | **11** |
| Female sex | -3 | -1 | -1 |
| Age | -1 | 0 | 0 |
| Hay fever | -1 | -1 | 0 |
| Pollen-triggered wheeze | -5 | -3 | -2 |
| Animal-triggered wheeze | -4 | -2 | -1 |
| Wheeze frequency 1 (1-3) | -2 | -1 | -1 |
| Wheeze frequency 2 (≥ 4) | -3 | -2 | -1 |
| Exercise-induced wheeze | -2 | -1 | -1 |
| Night-time awakening by wheeze | -8 | -4 | -2 |
| Paternal asthma | -2 | -1 | -1 |
| Maternal asthma | -2 | -1 | -1 |

**Table E8:** Development of the School-age Asthma Prognosis Score (SAPS): Performance comparison of candidate point-based models for asthma remission across different scaling factors

| | Score performance | | |
|---|---|---|---|
| **Scale** (x) | **Brier score** | **AUC** | **HL p-value** |
| 10 | 0.137 | 0.705 | 0.735 |
| 5 | 0.138 | 0.703 | 0.786 |
| 3 | 0.138 | 0.701 | 0.679 |

HL p-value after re-calibration.

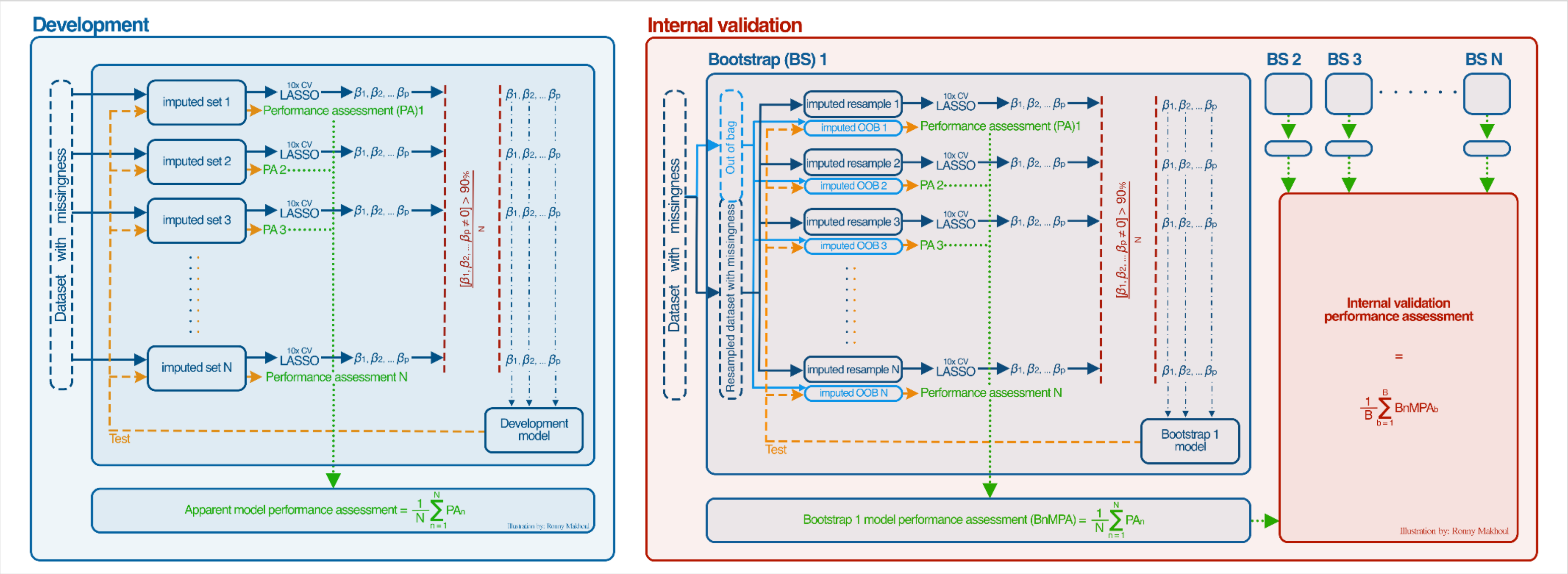

**Figure E1:** Model development and internal validation framework for the School-age Asthma Prognosis Score (SAPS).

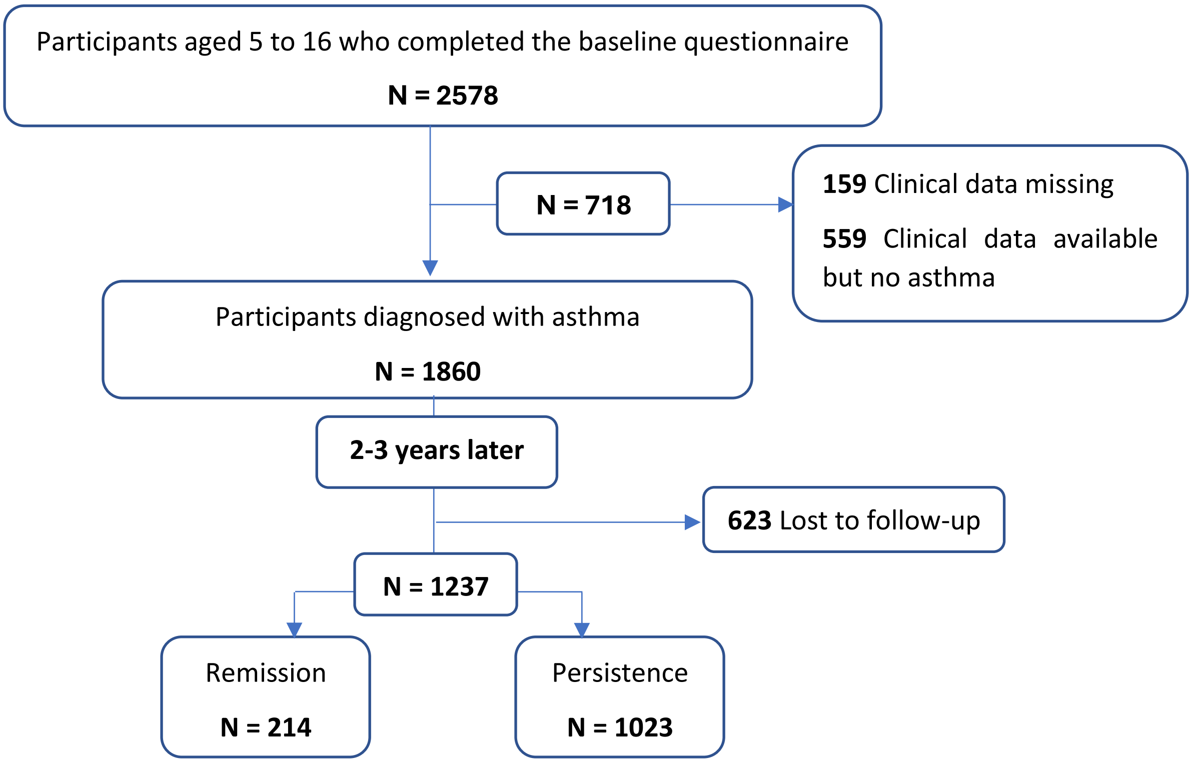

**Figure E2:** Flow diagram of participant inclusion for the School-age Asthma Prognosis Score (SAPS) development in the Swiss Paediatric Airway Cohort (SPAC).

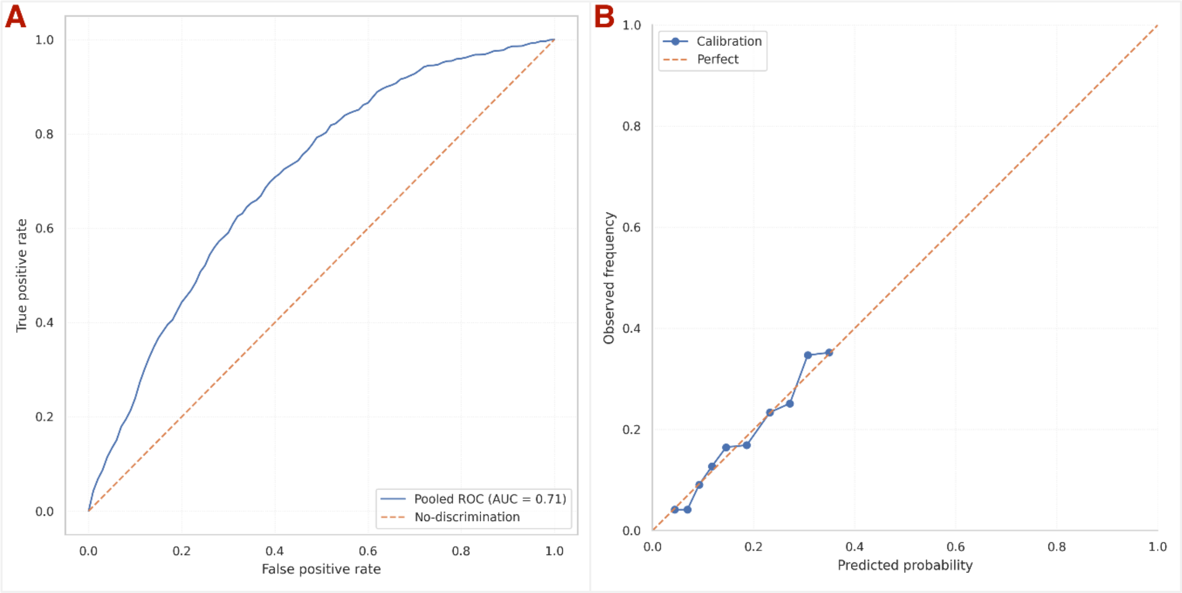

**Figure E3:** The School-age Asthma Prognosis Score (SAPS): Development model pooled ROC curve (A) and calibration plot (B).

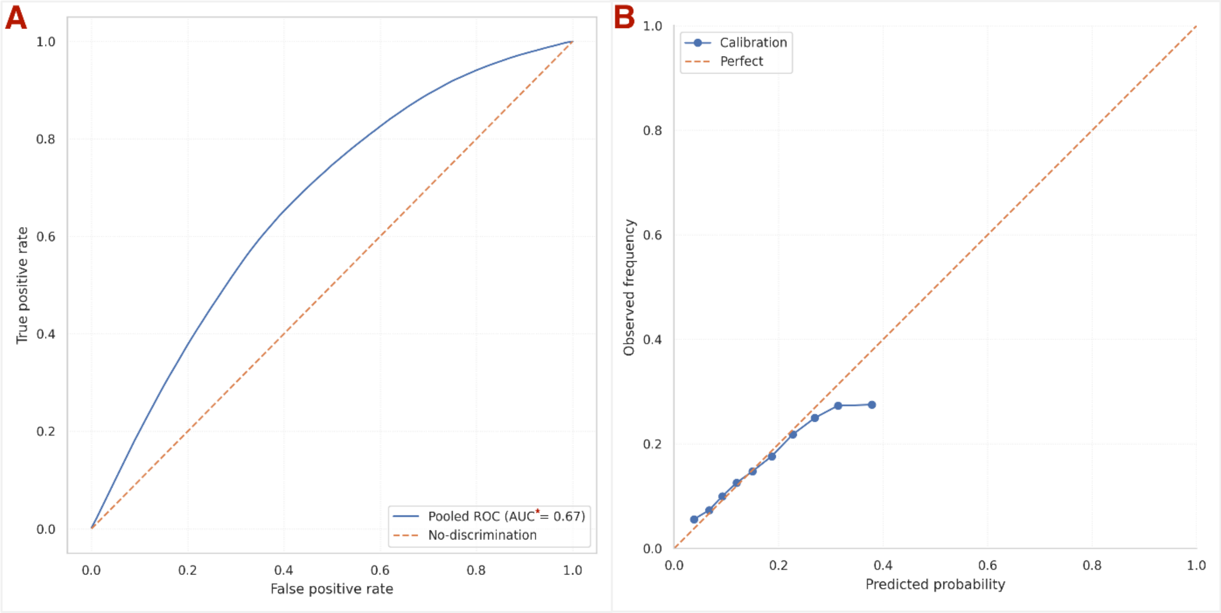

**Figure E4:** The School-age Asthma Prognosis Score (SAPS): Internal validation: pooled ROC curve (A) and calibration plot (B).

*Out-of-bag (OOB) AUC; optimism-corrected estimate reported separately.

**Supplementary Methods**

*Data examination and preprocessing*

Before model fitting, we examined the distributions and missingness patterns of all candidate predictors, cross-tabulated key variables to identify inconsistencies or unexpected associations, and compared baseline characteristics of participants with follow-up available to those lost to follow-up. We evaluated data completeness, and used multiple correspondence analysis to explore associations and potential redundancy among categorical variables.

*Handling of missing data in cases where we recoded missingness to zero*

This coding approach is based on the rationale that, in the SPAC study, parents and children are more likely to report symptoms and medication use when present, and non-response usually reflects absence, or ambiguous symptoms. This strategy has been recommended and used in previous cohort studies of asthma and respiratory symptoms, and reduces the risk of introducing bias due to unnecessary exclusions (E1-5).

*Sample and parameter calculation*

We determined sample size and model complexity in line with established recommendations for clinical prediction models, including guidance by Steyerberg and Riley (E6, E7). In the development cohort, 17% of children (214 of 1237) had asthma remission at follow-up. Using established rules, such as a minimum of 20 events per predictor variable (EPV), where a parameter is defined as each coefficient estimated in the model, including those for categorical predictors, and a multistep procedure from the pmsampsize R package (which are more conservative than EPV alone), the development (SPAC) dataset sample was sufficient to support development of a model with up to 13 parameters while minimizing the risk of overfitting. Specifically, a binary variable counts as one parameter, while a categorical variable with *k* levels contributes *k* – 1 parameters to the model (E6). We performed the calculations prior to multiple imputations.

*Development and apparent performance*

We compared baseline characteristics between participants with observed follow-up and those lost to follow-up and observed differences for several variables. This makes a missing completely at random (MCAR) mechanism unlikely. We therefore proceeded under a MAR assumption: conditional on observed data, loss to follow-up does not depend on the unobserved outcome. We used multiple imputation (m = 50) to address missingness in potential predictors, specifying imputation models that included the outcome and relevant auxiliary variables (E5, E8). Within each imputed dataset, we applied LASSO logistic regression, using ten-fold cross-validation to select the optimal penalty parameter. We retained predictors with nonzero coefficients in at least 90% of the imputed datasets. Then, for each predictor, we calculated the arithmetic mean of its coefficients across all imputations to create the development model. To assess apparent performance, we tested the development model on each of the imputed samples. We pooled – by calculating the arithmetic mean – performance results across all imputations to summarize model discrimination using the area under the receiver operating characteristic curve (AUC), calibration by calibration plots and the Hosmer-Lemeshow test, and overall accuracy with the Brier score. We generated visual summaries (pooled ROC and calibration plots) (E6).

*Internal validation*

For internal validation, we generated 600 bootstrap resamples from the original dataset. In each bootstrap replicate, we imputed the training resample and its out-of-bag (OOB) test set separately, generating 50 imputations for each, with outcome included in the imputation models for both (E8, E9). We fitted LASSO logistic regression on the imputed training resample sets and pooled the coefficients, following the same procedure used in the development phase. To evaluate model performance, we applied the pooled model to each OOB-imputed dataset. We pooled performance results within each bootstrap replicate across its OOB imputations and then aggregated the replicate-level estimates across all 600 bootstraps to obtain overall performance (**Supplementary Figure E1**). To correct for optimism, we combined this OOB estimate with the apparent performance from the development data using the Efron-Tibshirani 0.632+ estimator (E9, E10), computed for the AUC and the Brier score. Calibration was summarized directly from the OOB sets.

*Selection stability*

We quantified variable-selection stability from development to internal validation by recording selection frequencies for each predictor across imputations and bootstrap replicates and by summarizing agreement between selected sets using visual displays.

E4. Buuren Sv. Flexible Imputation of Missing Data. Second Edition ed. New York: Chapman and Hall/CRC; 2018.

E5. Carpenter JR, K. MG, Bartlett JW, Morris TP, Quartagno M, Wood AM. Multiple Imputation and its Application 2e. 2 ed: John Wiley & Sons, Ltd; 2023.

E6. Steyerberg EW. Clinical prediction models: A Practical Approach to Development, Validation, and Updating: Springer Cham; 2019.

E7. Riley RD, Ensor J, Snell KIE, Harrell FE, Jr., Martin GP, Reitsma JB, et al. Calculating the sample size required for developing a clinical prediction model. Bmj. 2020;368:m441.

E8. Moons KGM, Donders RART, Stijnen T, Harrell FE, Jr. Using the outcome for imputation of missing predictor values was preferred. Journal of Clinical Epidemiology. 2006(59, 1092-1101).

E9. Wahl S, Boulesteix AL, Zierer A, Thorand B, van de Wiel MA. Assessment of predictive performance in incomplete data by combining internal validation and multiple imputation. BMC Med Res Methodol. 2016;16(1):144.

E10. Efron B, Tibshirani R. Improvements on Cross-Validation: The .632+ Bootstrap Method.

| Section/Topic | Item | | Checklist Item | Page |
|---|---|---|---|---|
| **Title and abstract** | | | | |
| Title | 1 | D;V | Identify the study as developing and/or validating a multivariable prediction model, the target population, and the outcome to be predicted. | 1 |
| Abstract | 2 | D;V | Provide a summary of objectives, study design, setting, participants, sample size, predictors, outcome, statistical analysis, results, and conclusions. | 6 |
| **Introduction** | | | | |
| Background and objectives | 3a | D;V | Explain the medical context (including whether diagnostic or prognostic) and rationale for developing or validating the multivariable prediction model, including references to existing models. | 7 |
| | 3b | D;V | Specify the objectives, including whether the study describes the development or validation of the model or both. | 7 |
| **Methods** | | | | |
| Source of data | 4a | D;V | Describe the study design or source of data (e.g., randomized trial, cohort, or registry data), separately for the development and validation data sets, if applicable. | 8 |
| | 4b | D;V | Specify the key study dates, including start of accrual; end of accrual; and, if applicable, end of follow-up. | 8 |
| Participants | 5a | D;V | Specify key elements of the study setting (e.g., primary care, secondary care, general population) including number and location of centres. | 8 |
| | 5b | D;V | Describe eligibility criteria for participants. | 8 |
| | 5c | D;V | Give details of treatments received, if relevant. | n/a |
| Outcome | 6a | D;V | Clearly define the outcome that is predicted by the prediction model, including how and when assessed. | 9 |
| | 6b | D;V | Report any actions to blind assessment of the outcome to be predicted. | n/a |
| Predictors | 7a | D;V | Clearly define all predictors used in developing or validating the multivariable prediction model, including how and when they were measured. | 9 |
| | 7b | D;V | Report any actions to blind assessment of predictors for the outcome and other predictors. | n/a |
| Sample size | 8 | D;V | Explain how the study size was arrived at. | 10 |
| Missing data | 9 | D;V | Describe how missing data were handled (e.g., complete-case analysis, single imputation, multiple imputation) with details of any imputation method. | 10 |
| Statistical analysis methods | 10a | D | Describe how predictors were handled in the analyses. | 10-11 |
| | 10b | D | Specify type of model, all model-building procedures (including any predictor selection), and method for internal validation. | 10-11 |
| | 10c | V | For validation, describe how the predictions were calculated. | 10-11 |
| | 10d | D;V | Specify all measures used to assess model performance and, if relevant, to compare multiple models. | 10-11 |
| | 10e | V | Describe any model updating (e.g., recalibration) arising from the validation, if done. | n/a |
| Risk groups | 11 | D;V | Provide details on how risk groups were created, if done. | n/a |
| Development vs. validation | 12 | V | For validation, identify any differences from the development data in setting, eligibility criteria, outcome, and predictors. | 11 |
| **Results** | | | | |
| Participants | 13a | D;V | Describe the flow of participants through the study, including the number of participants with and without the outcome and, if applicable, a summary of the follow-up time. A diagram may be helpful. | 12 |
| | 13b | D;V | Describe the characteristics of the participants (basic demographics, clinical features, available predictors), including the number of participants with missing data for predictors and outcome. | 12 |
| | 13c | V | For validation, show a comparison with the development data of the distribution of important variables (demographics, predictors and outcome). | 12 |
| Model development | 14a | D | Specify the number of participants and outcome events in each analysis. | 12 |
| | 14b | D | If done, report the unadjusted association between each candidate predictor and outcome. | n/a |
| Model specification | 15a | D | Present the full prediction model to allow predictions for individuals (i.e., all regression coefficients, and model intercept or baseline survival at a given time point). | 12-13 |
| | 15b | D | Explain how to the use the prediction model. | 13 |
| Model performance | 16 | D;V | Report performance measures (with CIs) for the prediction model. | 12-13 (no CI) |
| Model-updating | 17 | V | If done, report the results from any model updating (i.e., model specification, model performance). | n/a |
| **Discussion** | | | | |
| Limitations | 18 | D;V | Discuss any limitations of the study (such as nonrepresentative sample, few events per predictor, missing data). | 15-16 |
| Interpretation | 19a | V | For validation, discuss the results with reference to performance in the development data, and any other validation data. | 14-15 |
| | 19b | D;V | Give an overall interpretation of the results, considering objectives, limitations, results from similar studies, and other relevant evidence. | 14-15 |
| Implications | 20 | D;V | Discuss the potential clinical use of the model and implications for future research. | 14-16 |
| **Other information** | | | | |
| Supplementary information | 21 | D;V | Provide information about the availability of supplementary resources, such as study protocol, Web calculator, and data sets. | Yes |
| Funding | 22 | D;V | Give the source of funding and the role of the funders for the present study. | 2 |

*Items relevant only to the development of a prediction model are denoted by D, items relating solely to a validation of a prediction model are denoted by V, and items relating to both are denoted D;V. We recommend using the TRIPOD Checklist in conjunction with the TRIPOD Explanation and Elaboration document. n/a: not applicable.
